# Association Between HIV PrEP Indications and Use in a National Sexual Network Study of Men Who Have Sex with Men

**DOI:** 10.1101/2021.03.03.21252823

**Authors:** Kevin M. Weiss, Pragati Prasad, Travis Sanchez, Steven M. Goodreau, Samuel M. Jenness

**Author notes:** Corresponding Author: Emory University, 1520 Clifton Road, Atlanta, GA, U.S.A. 30322.

## Abstract

**Background:** HIV preexposure prophylaxis (PrEP) is effective in preventing HIV transmission. US Public Health Service (USPHS) clinical practice guidelines define biobehavioral indications for initiation. To assess guideline implementation, it is critical to quantify PrEP non-users who are indicated and PrEP users who are not indicated.

**Methods:** Using data from a national web-based study of men who have sex with men (MSM) between 2017 and 2019, we estimated the association between PrEP use and PrEP indications. Log-binomial regression was used to estimate the relationship between PrEP indications and PrEP use, adjusted for geography and demographics.

**Results:** Of 3508 sexually active, HIV-negative MSM, 34% met indications for PrEP. The proportion with current PrEP use was 32% among those meeting indications and 11% among those without indications. Nearly 40% of those currently using PrEP did not meet indications for PrEP, and 68% of MSM with indications for PrEP were not currently using PrEP. After adjusting for geography and demographics, MSM with PrEP indications were about 3 times as likely to be currently using PrEP. This association varied slightly by geography and demography.

**Conclusions:** Indications for PrEP strongly predicted current PrEP use among MSM. However, we identified substantial misalignment between indications and use in both directions (indicated MSM who were not benefitting from PrEP, and MSM taking PrEP while not presently being indicated). This calls for further implementation efforts to improve PrEP delivery to those most in need during periods of elevated sexual risk.

## INTRODUCTION

Despite biomedical advances in HIV prevention with antiretroviral pre-exposure prophylaxis (PrEP), the burden of HIV among men who have sex with men (MSM) remains high.^1^ MSM are a high-priority risk group for PrEP use based on their behavioral and biological risk factors.^2,3^ The US Public Health Service (USPHS) produced clinical provider guidelines, most recently updated in 2017, which define specific bio-behavioral indications for PrEP prescription for MSM.^4^

The population impact of PrEP depends on coverage, or the proportion of indicated persons who use PrEP. Mathematical models and ecologic implementation data have estimated the association between PrEP coverage and lower HIV incidence^5^ and diagnosis rates among MSM.^6^ Data through the mid-2010s indicated that only a small fraction of sexually active MSM were estimated to be using PrEP^7–9^, though uptake has increased over the past few years, with two recent estimates placing uptake among eligible MSM at 20%^10^ and 35%.^11^ Various factors such as access to health care^12^ likely contribute to suboptimal PrEP coverage. There is some evidence that patient-perceived HIV risk may underestimate clinical assessments of HIV risk and eligibility for PrEP.^13^ It is critical to characterize the group of MSM with indications for PrEP but who are not using it.

To achieve maximum prevention benefits under financial constraints, PrEP implementation efforts must also consider efficiency. Efficient delivery of PrEP means a low number needed to treat (NNT, quantified as person-time on PrEP) to avert one new infection. Maximally efficient intervention targeting scenarios, which deliver interventions only to a group who would optimally benefit from it, have an NNT approaching 1. PrEP use in some groups of MSM who are at low likelihood of acquiring HIV (either through their own behavior or as a function of their epidemiological context) could substantially reduce PrEP efficiency while having a minimal impact on HIV incidence.^5^ PrEP medication supplies are not currently limited and care rationing is unlikely, yet use of PrEP clinical services by MSM not indicated for PrEP may also limit the potential efficacy of PrEP implementation.

In this study, we explore the two “off-diagonal” scenarios in the PrEP indication and uptake matrix: those who are indicated for PrEP but not using it, and those who are not indicated for PrEP but using it. Although the indications in the guidelines have imperfect sensitivity and specificity in capturing MSM who may be at risk for HIV acquisition, examining this misalignment may be the first step towards their reevaluation. It is particularly important to determine whether these associations differ among key MSM subgroups commonly focused on in HIV prevention research to understand how misalignment may contribute to or correlate with differential HIV incidence and PrEP use.

Our primary research aims were to estimate the proportions of MSM ever or currently using PrEP compared to their indications, and to characterize the predictors of misalignment. We hypothesized that PrEP indications would strongly predict PrEP use, but that there would be misalignment of these proportions and variations by geography and race/ethnicity.

## METHODS

### Study Design

We used data from the ARTnet study of cis-gender MSM in the United States for this analysis. The complete methods for ARTnet have been described previously.^14,15^ ARTnet is a web-based sexual network study to characterize the sexual partnership networks and engagement in HIV prevention services of MSM in the United States. Eligibility included any lifetime history of male-male sex and age between 15 and 65 years. ARTnet participant data were linked to the participant’s responses from the American Men’s Internet Study^16^ and then de-duplicated. Study procedures were completed between 2017 and 2019. The Emory University Institutional Review Board approved the study protocol. The main study procedure was an online survey, which was hosted on a HIPAA-compliant platform (SurveyGizmo, Boulder, CO).

### PrEP Use

Two measures were used to calculate PrEP use. All participants reporting a negative result on their last HIV test were provided with a short description of PrEP as an antiretroviral pill (Truvada), which could be taken every day to reduce a person’s chance of getting HIV. Participant lifetime use of PrEP was then measured by the question “Have you ever taken PrEP (i.e. Truvada)?” with participants who responded affirmatively also having current PrEP uptake assessed by the question: “Are you currently taking PrEP (i.e. Truvada)?”

### PrEP Indications and Covariates

Survey data were used to evaluate whether participants reported behavior that defined indications under the updated USPHS clinical provider practice guidelines for PrEP prescription.^4^ These measures included the number of recent sexual partners and, additional individual- and partnership-level information. for up to the five most recent partners reported in the year prior to survey completion. These measure included: whether the partnership was with a main, casual, or one-time partner, the dates of the partnership (start, end, most recent sexual activity, whether the participant thought that the relationship would continue), what sexual activities occurred with each partner (anal/oral intercourse, frequency of sexual acts, sexual role and positioning), whether the participant and partner were using PrEP or antiretroviral therapy or condoms, and the respondent and partner’s histories of STI diagnosis.

Two sub-populations were defined for the analysis. The first, referred to as the *PrEP-eligible population*, includes all MSM who reported being HIV-negative and having been sexually active with another man in the prior 12 months. For the second, the subset of MSM meeting all USPHS indications for PrEP^4^ was then defined, referred to hereafter as *PrEP-indicated*. To be categorized as indicated, participants had to: 1) be aged 18 or older (excluded for 15–17 year old age category to evaluate eligibility based on behavior); 2) have their most recent HIV test be negative 3) not be in a monogamous partnership (per respondent definition) with an HIV-negative partner; 4) report anal intercourse with another man in the prior 6 months; and 5) report either any condomless anal intercourse with another man in the prior 6 months or a diagnosis of gonorrhea, chlamydia, or syphilis in the prior 6 months.

Participant-reported survey data were collected for other covariates. Participant-reported ZIP Codes were matched to one of four census regions (South, Midwest, Northeast, West),^17^ while reported race/ethnicity were grouped into four categories: Non-Hispanic Black, Non-Hispanic White, Hispanic, Other race/ethnicity. Additionally, reported ages were grouped into categories: 15–17; 18–24; 25–34; 35– 44; 45–54; and 55–65.

### Statistical Analysis

For our exploratory analysis, we present descriptive analyses of persons indicated for and using PrEP by demographic category using proportions and standard deviations. To quantify the association between PrEP indications and use, we selected a primary exposure of having USPHS indications for PrEP and outcome of current PrEP use. We used log-binomial regression models to quantify this association with prevalence ratios. After calculating the crude association, we estimated the multivariable relative risks after adjustment for race/ethnicity and geography. We also explored these covariates in an interaction model in order to evaluate how the association between PrEP indications and current PrEP use differed by demographics. All data analysis was performed in *R* 3.4.^18^

## RESULTS

Table 1 summarizes the interactions between indications for PrEP and PrEP use. Among 4904 study participants, 3508 PrEP-eligible men were identified. Overall, one-quarter (25.8%) had ever used PrEP, while slightly fewer (18.0%) were currently using PrEP. More than one-third (34.0%) met indications for PrEP. Among those meeting indications for PrEP, 41.0% reported ever using PrEP while 32.2% reported current PrEP use. Of men who did not meet indications for PrEP, lifetime and current PrEP use were 17.4% and 10.6%. More than one-half (56.3%) of those who had ever taken PrEP currently met indications for PrEP, while a slightly larger proportion (61.0%) of those currently taking PrEP met indications for PrEP. Thus, 39.0% of current PrEP users did not meet indications for PrEP, while two-thirds (67.8%) of PrEP-indicated MSM were not currently using PrEP. In total, 7.0% (246) of PrEP-eligible MSM were currently using PrEP despite not meeting indications for PrEP, while 23.1% (809) of PrEP-eligible MSM were not using PrEP despite meeting indications for PrEP.

**Table 1.**
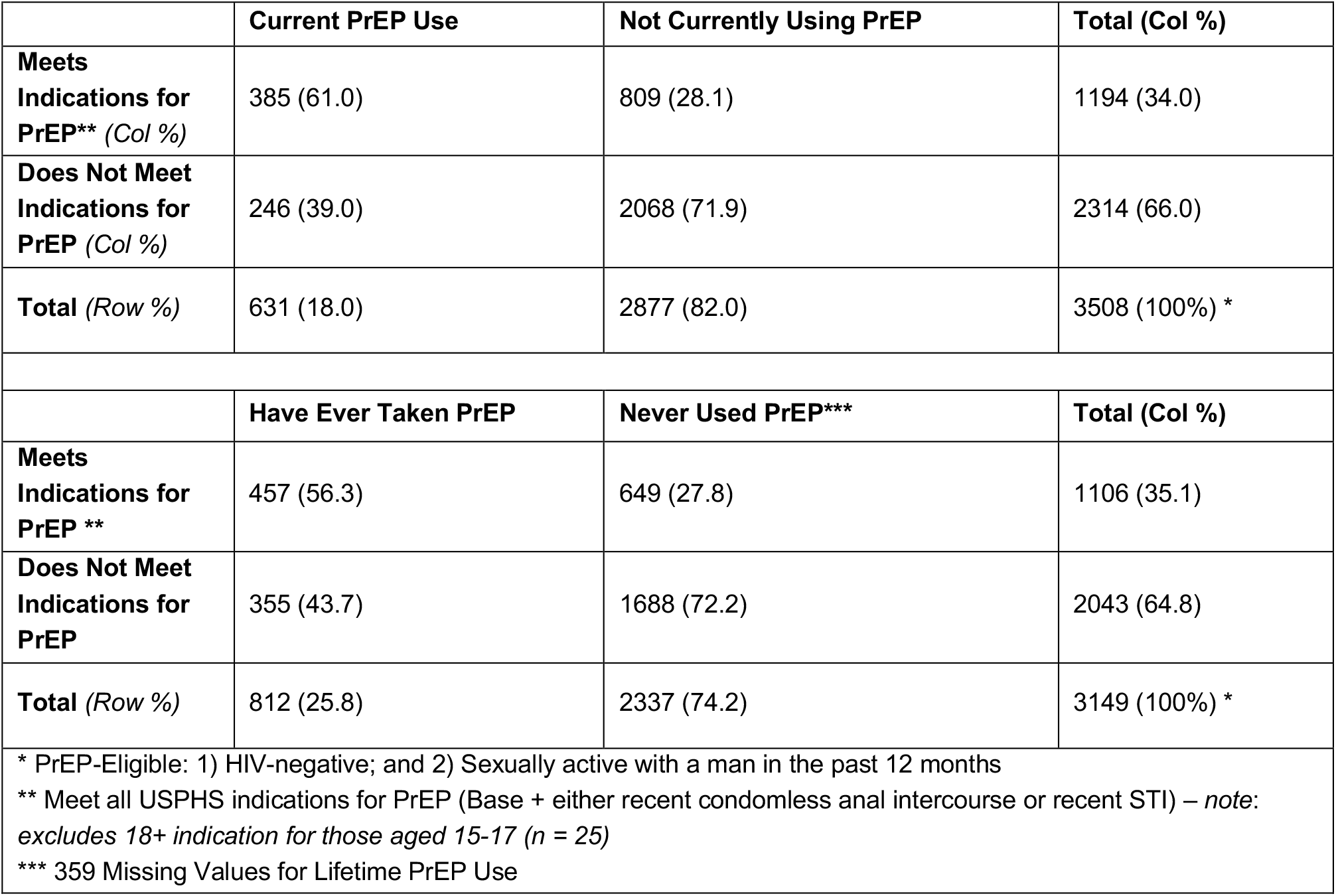
Indications for PrEP and Current and Ever PrEP Use among PrEP-Eligible MSM*

|  |  |  |  |
| --- | --- | --- | --- |
|  | **Current PrEP Use** | **Not Currently Using PrEP** | **Total (Col %)** |
| **Meets Indications for PrEP\*\* (Col %)** | 385 (61.0) | 809 (28.1) | 1194 (34.0) |
| **Does Not Meet Indications for PrEP (Col %)** | 246 (39.0) | 2068 (71.9) | 2314 (66.0) |
| **Total (Row %)** | 631 (18.0) | 2877 (82.0) | 3508 (100%) * |
|  | **Have Ever Taken PrEP** | **Never Used PrEP\*\*\*** | **Total (Col %)** |
| **Meets Indications for PrEP \*\*** | 457 (56.3) | 649 (27.8) | 1106 (35.1) |
| **Does Not Meet Indications for PrEP** | 355 (43.7) | 1688 (72.2) | 2043 (64.8) |
| **Total (Row %)** | 812 (25.8) | 2337 (74.2) | 3149 (100%) * |
| * PrEP-Eligible: 1) HIV-negative; and 2) Sexually active with a man in the past 12 months<br>** Meet all USPHS indications for PrEP (Base + either recent condomless anal intercourse or recent STI) – *note: excludes 18+ indication for those aged 15-17 (n = 25)*<br>*** 359 Missing Values for Lifetime PrEP Use |  |  |  |

Table 2 summarizes current PrEP usage by demographic characteristics and PrEP indication status. The percentage of PrEP-indicated MSM who were currently using PrEP varied between 14.1% and 38.4% (32.2% overall). Among MSM meeting indications for PrEP, there was little variation by race/ethnicity. Geographically, current PrEP use among indicated MSM was lowest in the South (28.7%) and greatest in the West (38.4%). PrEP use increased with age, with lowest use among 15–17 (16.7%) and 18-24-year-old MSM (14.1%) and increasing use in age groups greater than 24 years, culminating in highest PrEP use (38.1%) among 45-54-year-old MSM. Across all demographic and geographic subgroups, there was a sizeable pool of PrEP-eligible MSM who met indications for, but were not currently taking, PrEP. Young (<25), Hispanic, and Western or Southern MSM comprised the greatest proportions of indicated men not currently using PrEP. Overall, 1 in 10 (10.6%) MSM who did not meet indications were currently taking PrEP, with use most likely among middle-aged MSM (35-54) and least likely among Hispanic, Southern, and young (<25) MSM.

**Table 2.** PrEP Use by Indication Status and Demographics among PrEP-Eligible MSM*

|  | MSM Meeting Indications for PrEP** |  |  | MSM Not Meeting Indications for PrEP** |  |  | Current PrEP Users |
| --- | --- | --- | --- | --- | --- | --- | --- |
|  | Total | Current PrEP Use | Not Using PrEP | Total | Current PrEP Use | Not Using PrEP | Meet Indication |
|  | *N* | *N (Row %)* | *N (Row %)* | *N* | *N (Row %)* | *N (Row %)* | % |
| **Overall** | 1194 | 385 (32.2) | 809 (67.8) | 2314 | 246 (10.6) | 2068 (89.4) | 61.0 |
| ***Race/Ethnicity*** |  |  |  |  |  |  |  |
| Non-Hispanic White | 871 | 284 (32.6) | 587 (67.4) | 1705 | 189 (11.1) | 1516 (88.9) | 60.0 |
| Non-Hispanic Black | 49 | 16 (32.7) | 33 (67.3) | 104 | 10 (9.6) | 94 (90.4) | 61.5 |
| Hispanic | 172 | 52 (30.2) | 120 (69.8) | 302 | 27 (8.9) | 275 (91.1) | 65.8 |
| Other | 102 | 33 (32.4) | 69 (67.6) | 203 | 20 (9.9) | 183 (90.1) | 62.3 |
| ***US Census Region*** |  |  |  |  |  |  |  |
| Northeast | 214 | 71 (33.2) | 143 (66.8) | 436 | 51 (11.7) | 385 (88.3) | 58.2 |
| Midwest | 252 | 75 (29.8) | 177 (70.2) | 440 | 51 (11.6) | 389 (88.4) | 59.5 |
| South | 418 | 120 (28.7) | 298 (71.3) | 853 | 81 (9.5) | 772 (90.5) | 59.7 |
| West | 310 | 119 (38.4) | 191 (61.6) | 585 | 63 (10.8) | 522 (89.2) | 65.4 |
| ***Age Category*** |  |  |  |  |  |  |  |
| 15–17 | 6 | 1 (16.7) | 5 (83.3) | 19 | 0 (0.0) | 19 (100.0) | 100.0 |
| 18–24 | 213 | 30 (14.1) | 183 (85.9) | 551 | 40 (7.3) | 511 (92.7) | 42.9 |
| 25–34 | 331 | 118 (35.6) | 213 (64.4) | 693 | 81 (11.7) | 612 (88.3) | 59.3 |
| 35–44 | 205 | 76 (37.1) | 129 (62.9) | 342 | 44 (12.9) | 298 (87.1) | 63.3 |
| 45–54 | 252 | 96 (38.1) | 156 (61.9) | 349 | 52 (14.9) | 297 (85.1) | 64.9 |
| 55–65 | 187 | 64 (34.2) | 123 (65.8) | 360 | 29 (8.1) | 331 (91.9) | 68.8 |

\* PrEP-Eligible: 1) HIV-negative; and 2) Sexually active with a man in the past 12 months
\*\* Meet all USPHS indications for PrEP (Base + either recent condomless anal intercourse or recent STI) - *note: excludes 18+ indication for those aged 15-17 (n = 25)*

Table 2 also includes the proportion meeting PrEP indications among MSM using PrEP. Variation in meeting PrEP indications varied little by race/ethnicity, ranging from 60.0% among Non-Hispanic white MSM to 65.8% of Hispanic MSM. PrEP-using MSM in the West (65.4%) were the only geographic subgroup where the percentage meeting indications for PrEP exceeded the overall average (61.0%). Younger PrEP-using MSM were less likely to meet indications for PrEP, with the notable exception of the small 15-17-year-old cohort, with proportions increasing by age.

In crude analyses, PrEP-eligible men who met indications for PrEP were 3.03 (95% Confidence Interval: 2.63, 3.51) times as likely as those not meeting indications to be currently taking PrEP (**Table 3**). In bivariate analyses, men living in the Northeast and West were 1.19 (0.97, 1.45) and 1.29 (CI: 1.07, 1.54) times, respectively, as likely as men in the South to be currently using PrEP. After adjusting for geography, age, and race/ethnicity, there was little change in the magnitude of the association (prevalence ratio (PR) = 2.98). There was a slightly higher probability of current PrEP use with increasing age. The relative risk of current PrEP use among non-white participants was marginally lower (from 5% to 9% lower) than white participants in crude analyses.

**Table 3.** Crude and Adjusted Log-Binomial Regression Correlates of Current PrEP use among PrEP-Eligible MSM

| Correlate | Crude Prevalence Ratio (95% CI) | Adjusted Prevalence Ratio (95% CI) * |
| --- | --- | --- |
| ***PrEP Indications*** |  |  |
| Meet Indications for PrEP | 3.03 (2.63, 3.51) | 2.98 (2.59, 3.45) |
| ***Census Region*** |  |  |
| Northeast | 1.19 (0.97, 1.45) | 1.18 (0.97, 1.43) |
| Midwest | 1.15 (0.94, 1.41) | 1.10 (0.90, 1.33) |
| South | --- | --- |
| West | 1.29 (1.07, 1.54) | 1.30 (1.09, 1.55) |
| ***Age*** |  |  |
| Age (10 years) | 1.12 (1.07, 1.18) | 1.10 (1.04, 1.15) |
| ***Race/Ethnicity*** |  |  |
| Non-Hispanic White | --- | --- |
| Non-Hispanic Black | 0.93 (0.63, 1.29) | 1.02 (0.70, 1.40) |
| Hispanic | 0.91 (0.72, 1.12) | 0.93 (0.97, 1.15) |
| Other | 0.95 (0.72, 1.21) | 0.98 (0.75, 1.24) |

The association between meeting PrEP indications for PrEP and using PrEP varied by subgroup (**Table 4**). The association was strongest among MSM in the West (PR: 3.56) and weakest in the Midwest (PR: 2.57). When evaluating the relationship among racial/ethnic participant subgroups, the magnitude of association was strongest among non-Hispanic black (PR: 3.40) and Hispanic (PR: 3.38) participants and weakest among white participants (PR: 2.94).

**Table 4.** Prevalence Ratio for Association between PrEP Indications and PrEP Use in Subgroup Models among PrEP-Eligible MSM

| Correlate | Prevalence Ratio (95% CI) |
| --- | --- |
| ***Census Region*** |  |
| Northeast | 2.84 (2.07, 3.93) |
| Midwest | 2.57 (1.87, 3.56) |
| South | 3.02 (2.35, 3.92) |
| West | 3.56 (2.73, 4.71) |
| ***Race/Ethnicity*** |  |
| Non-Hispanic White | 2.94 (2.50, 3.47) |
| Non-Hispanic Black | 3.40 (1.69, 7.24) |
| Hispanic | 3.38 (2.23, 5.26) |
| Other | 3.28 (2.01, 5.54) |

## DISCUSSION

In this study, we found evidence of substantial misalignment between the USPHS indications for PrEP and current use of PrEP. The misalignment was greater for MSM indicated for PrEP but not using it compared to MSM not indicated for PrEP but using it. Overall, 68% of PrEP-indicated MSM (23% of all PrEP-eligible MSM) were not using PrEP, and 39% of MSM currently using PrEP (7% of all PrEP-eligible MSM) were not indicated for PrEP. Having indications for PrEP strongly predicted PrEP use, but nearly one-third of PrEP-eligible MSM had possible misalignment between indications and use. This suggests overall that PrEP underuse remains a public health priority nationally, with continued efforts needed to close the gap between indications and uptake.

Control of the HIV epidemic with PrEP depends on a relatively high level of coverage,^5,6^ but persistent PrEP underuse among at-risk MSM limits the possibility of meeting federal and local HIV prevention goals.^19^ We found that a significant number of MSM with biobehavioral indications, indicating increased risk for HIV acquisition, such as a recent STI or recent CAI were not using PrEP, aligning with previous work focusing on the interaction of eligibility and use.^20^ Using the framework of a PrEP care continuum,^21,22^ many individual- and structural-level barriers may limit otherwise suitable candidates from benefitting from using the medication. This includes decreased HIV risk perception,^13^ which may be driven by suboptimal provider and patient knowledge of the USPHS provider guidelines. These barriers, however, highlight potential points of interventions to improve access, use, adherence, and persistence on PrEP.^8,23,24^

Although less prevalent than underuse, PrEP use by those not meeting indications could potentially also impede efficient PrEP implementation. Previous modeling has shown that the efficiency of PrEP (e.g., number needed to treat) for both HIV and associated prevention depends on the target population, with decreased efficiency when PrEP is provided to individuals who are at lower risk.^5,25^ Theoretically, though never observed, use of PrEP by those without behavioral indications, or a “worried well” population, requires societal resources (in terms of public and private funding of medications via health care insurance payments) and use of clinical services that may not generate a large clinical benefit. To the hypothesis that provider and patient knowledge of PrEP and HIV risk can be improved, the guidelines for PrEP indications provide a benchmark for how PrEP determination can be assessed.^26^ However, this relies on ensuring trust between patients and providers to obtain an accurate sexual history.^24^ As PrEP scale-up continues, it is essential that resources be given to those who may benefit the most from PrEP, including associated services such as regular STI screening, and may face the greatest barriers to uptake.

Few other assessments have able to concurrently present PrEP eligibility alongside PrEP use, as this ARTnet estimate does.^10,11,27^ When compared to these studies, PrEP uptake estimates for MSM in this online study (18.0%) were slightly lower. For example, approximately one-quarter of HIV-negative MSM in major metropolitan areas surveyed through National HIV Behavioral Surveillance among MSM reported using PrEP in the past 12 months, which tracked closely with ARTnet estimates of lifetime PrEP usage (not presented) and exceeded current PrEP usage in ARTnet.^11^ These differences may reflect differences in study data collection (online vs. in-person) as well as different sampling frames (e.g. urban MSM). Given the ubiquity of internet access among US residents^28^ and the experiences of large internet-based surveys of sexual behaviors among MSM,^29^ web-based studies can provide a feasible, cost-effective estimates of PrEP indications and uptake among MSM to complement database-driven estimates, though further work to assess why these estimates may differ is warranted.

Given the observed gaps in both HIV incidence and PrEP use among MSM of color, particularly black and Hispanic MSM, the association between PrEP indications and PrEP use matters for disparity reduction. Previous research has demonstrated that persons of color have an equal or greater proportion with indications for PrEP,^7^ but make up only a small fraction of PrEP prescriptions.^30^ MSM of color have seen HIV diagnosis rates remain stable or increase while rates decreased among white MSM,^1^ have lower comparative levels of PrEP use,^27^ and, in some studies, persons of color were less likely to be indicated for or receive PrEP.^31^ In our analysis, the magnitude of association between PrEP indications and current PrEP use was greatest among non-Hispanic black and Hispanic participants and least among non-Hispanic white participants. This finding does not necessarily align with studies that have found difficulty translating interest into uptake^32^ and a lagged awareness of PrEP and HIV prevention in these same populations.^33^ Multiple explanations are possible and likely contribute to this context, including potential individual (behavioral, psychological, risk perception, stigma, medical mistrust, self-efficacy)^13,23,34^ and structural (insurance, cost, access to care, health utilization) barriers to accessing and using PrEP for black and other MSM of color that may reduce uptake and lead to decreased sensitivity of the USPHS guidelines in assessing HIV acquisition risk.^35^

Given the high HIV burden and challenges with accessing healthcare in the US South, we hypothesized that PrEP use there would be lower than in other regions. The highest rates of HIV diagnosis are in the US South,^1^ as well as the lowest levels of PrEP use.^36–38^ PrEP clinics are unevenly distributed across the US,^39^ and the current number of clinics in the South may not be able to support the estimated need for PrEP.^40^ In this study, surveyed MSM in the US South indeed reported the lowest levels of current PrEP usage among the four census regions, though the pattern was not dissimilar from other regions such as the Midwest. Additionally, even among MSM meeting indications for PrEP, MSM from the South were least likely to be taking PrEP. As a crude measure of regional differences, the magnitude of the association between PrEP indications and use varied more geographically than when stratified by other characteristics, although it was not weakest in the South. These crude measures may be influenced by other variables (possibly race/ethnicity), and likely need further exploration to determine their importance and whether they further highlight additional barriers to starting PrEP. A recent review of PrEP implementation strategies in the South highlighted a number of individual and structural factors, including a greater rural population, lesser access to PrEP care, fewer insured individuals, lower health literacy and HIV risk perception, and greater anti-HIV, anti-gay, and PrEP stigma, that likely influence PrEP uptake in the South.^41^ Novel implementation strategies, such as telemedicine-based PrEP to better reach people in rural areas^42,43^ or consideration of networks and social capital,^44,45^ may be necessary to try to offset the barriers observed for PrEP uptake in the South.

### Limitations

This analysis has some limitations. These data represent a convenience sample of MSM recruited online from across the US and are not representative of all MSM, including potential under-representation of racial and ethnic minority MSM.^46^ Ideally, these results could be interpreted in the context of other representative samples of MSM, but none exist. However, we conducted stratified analysis by three core factors (race, age, geography) in which there may be imbalances to estimate group-specific outcomes. With the assumption of conditional exchangeability, this partially alleviates this issue. In any online study, such as ARTnet, social desirability bias may be a factor, potentially resulting in overestimation of desirable prevention behaviors such as PrEP uptake. Additionally, to be asked questions about PrEP uptake, MSM had to disclose or report being HIV-negative, which could be subject to potential social desirability bias, thus overestimating the number of PrEP-eligible MSM, as well as being influenced by a reluctance to disclose personal information, which could have underestimated the number of PrEP-eligible MSM. Our use of most recent HIV result to determine HIV status could underestimate the proportion of MSM with indications for PrEP by undercounting MSM who are at truly at-risk (sexually active) but have not recently tested or ever have been tested.

## Conclusions

Routine monitoring of PrEP uptake is needed to measure progress towards and gaps in PrEP coverage. Lifetime and current PrEP uptake among MSM exceeded previous estimates, and meeting PrEP indications was strongly associated with current PrEP usage. However, there remain populations of MSM who are indicated but not using PrEP, as well as MSM who are using PrEP while not currently meeting the indications. The relative importance of behavioral indications and demographic differences for PrEP uptake, amidst recent events that aim to address structural barriers to accessing PrEP, including the US government’s Ending the Epidemic program^19^, Ready, Set, PrEP initiative,^47^ and the US Preventive Service Task Force recommendation requiring coverage without cost-sharing,^48,49^ highlight potential barriers to and gaps in implementing PrEP nationally which will need to be addressed to meet PrEP’s full potential to reduce new infections across the US.

## Data Availability

ARTnet data are available to external researchers upon completion of a data use agreement, given the sensitive nature of the study data and IRB restrictions.

